# A mixed-methods evaluation of patients’ views on primary care multi-disciplinary teams in Scotland

**DOI:** 10.1101/2023.08.29.23294713

**Authors:** KD Sweeney, E Donaghy, D Henderson, HHX Wang, SW Mercer

## Abstract

**Background:** Expanding the primary care multi-disciplinary team (MDT) is a key aim of the 2018 Scottish GP contract, and over 3,000 new MDT-staff have been appointed since then.

**Aim:** To explore patients’ views on MDT expansion in primary care in Scotland.

**Design and methods:** Survey of patients aged 18 years and over who had consulted a GP in the previous four weeks, in three population settings (deprived urban (DU), affluent urban (AU) and remote and rural (RR)), followed by 30 semi-structured individual interviews. The survey assessed awareness of five key new MDT roles, and attitudes towards reception signposting. Interviews explored views regarding MDT-care generally.

**Results:** Of the 1,053 survey respondents, most were unaware of the possibility of being offered MDT, rather than GP, consultations, for three out of five roles (69% unaware of link worker appointments; 68% mental health nurse; 58% pharmacist). Reception signposting to MDT was viewed significantly more negatively in DU areas than elsewhere (34% quite or very unhappy vs 21% AU vs 29% RR; p<0.001).

Most of the 30 patients interviewed were accepting of MDT-care, and many reported positive first-hand experiences. Improved access and added expertise were perceived benefits. However, many had reservations about MDT expansion and an overriding preference for holistic, relationship-based GP-care.

**Conclusion:** Four years since the introduction of the new Scottish GP contract, patient awareness of MDT expansion is limited, views on reception signposting mixed, though experiences of MDT-care generally positive. However, patients still want to see a known GP when they feel it is important, and report this as being challenging especially in deprived areas.

## Introduction

Multi-disciplinary teams (MDTs) are an increasingly common feature of primary care systems^1-5^. Services that bring together a range of healthcare professionals to deliver care alongside general practitioners (GPs) are assumed to be better equipped to provide accessible and comprehensive primary care^4-8^, especially given the global shortage of GPs^9, 10^. MDT-based models of care (also referred to as interdisciplinary care, interprofessional care, or interprofessional collaborative practice) have been shown in some studies to improve health outcomes and reduce healthcare costs^11, 12^. Accordingly, the expansion of MDTs has been a common feature of recent primary care reforms in high income countries, as health systems seek to address challenges of increasing multimorbidity, population ageing, growing health inequalities and rising costs associated with secondary care^13^.

In Scotland, the expansion of MDTs in primary care was a key part of the new GP contract introduced in April 2018 against a background of service redesign^14^. Healthcare delivery is organised within 14 regional Health Boards in Scotland. In April 2016, the Quality Outcome Framework pay-for-performance system was abolished and GP practices began to be organised into geographic Clusters to promote shared quality improvement and local integration of care. The 2018 GP contract formalised these changes, and further refocussed the role of the GP as the expert medical generalist at the heart of a multi-disciplinary team of professionals including practice nurses, advanced nurse practitioners, physiotherapists, mental health nurses, pharmacists and community link workers^14^. By increasing the amount of care provided by healthcare professionals other than GPs, the contract aimed to improve the quality and accessibility of care for patients, while reducing GP workload^14^. In turn, this was intended to enable GPs to focus more time on patients with complex needs. Between 2018 and 2022, 3,220 whole-time equivalent new MDT-staff were appointed in primary care in Scotland.

As well as addressing pressures of increasing multimorbidity and population ageing, a stated aim of the 2018 GP contract was the mitigation of health inequalities, which are widening in Scotland^15^. The expansion of primary care MDTs – including community link workers who focus on connecting patients to non-medical community resources in deprived areas– was central to this. Whether the new GP contract has effectively contributed to the mitigation of health inequalities is not known, but our recent survey of patient experiences and qualitative evaluations of the views of senior primary care stakeholders, GPs, MDT staff, and patients living in deprived areas suggest it has not^16-18^.

Most evaluations of MDT-based models of primary care have focussed on health outcomes and costs^11, 12^. Our aforementioned qualitative evaluations explored the views of a range of professionals with regard to the new GP contract in Scotland^16, 17^. However, the views and experiences of patients remains underexplored^8, 13, 19, 20^. This study aims to assess the awareness, attitudes and experiences of patients with respect to primary care MDT expansion in Scotland, following the introduction of the 2018 GP contract.

## Methods

### Study Design

The study consisted of a postal questionnaire distributed to patients who had recently consulted a GP, followed by individual telephone interviews with a subsample of respondents.

### Sampling and recruitment

Twelve GP practices were purposively sampled from three Health Boards to give a range of geographic and socioeconomic characteristics, as fully described in our previous paper^18^. A random sample of 6291 adult patients who had consulted a GP within the past 30 days were identified from practice records.

Surveys were sent with a cover letter which included an optional expression of interest form to identify potential participants for follow-up qualitative interviews. Surveys were posted between 31^st^ August and 15^th^ September. Collection of responses ran until 30^th^ November 2022. Interviews were conducted between 26^th^ October 2022 and 11^th^ January 2023.

1053 questionnaire responses were received (response rate 17%). Response rates were higher in AU areas (27%) than in RR areas (20%) and DU areas (12%). Of those who returned expression of interest forms, 30 patients were purposively sampled for interview (10 from each Health Board), with two-thirds aged over 65 years, two-thirds with multimorbidity (two or more chronic conditions).

### Data collection

The questionnaire collected sociodemographic information including respondents’ age, gender and deprivation status, which was obtained from the Scottish Index of Multiple Deprivation (SIMD) linked with each patient’s postcode, and recorded in deciles, with 1 being the most deprived 10% and 10 the least deprived 10%^21^. Multimorbidity was assessed using a checklist of 17 common chronic conditions, with space to add additional conditions not listed, as in our previous studies^22, 23^.

The questionnaire included several validated and bespoke items assessing health characteristics, patterns of consulting, and consultation experience, results from which have been published separately^18^. For the present paper, the questionnaire included bespoke items assessing respondents’ awareness (using binary response options “I am aware of this” or “I don’t know about this”) of the possibility of being offered an appointment with the following five MDT professionals: a nurse (including practice nurse, advanced nurse practitioner or health care assistant); a mental health nurse based at the practice; a physiotherapist; a pharmacist or pharmacy technician based at the practice; and a community link worker or welfare advisor. Respondents were also asked how happy they would feel (using a four-point Likert scale) if they were asked about their health concern by the receptionist for the purpose of signposting, if appropriate, to an MDT professional without seeing a GP first.

One-to-one semi-structured telephone interviews lasting 40-60 minutes were conducted with the 30 selected patients. Interviews were audio recorded and transcribed verbatim. The interview topic guide covered patients’ views and experiences regarding access to appointments, GP care, MDT care, telephone consultations and the impact of the Covid-19 pandemic. The present paper focusses on MDT-care and reception signposting, with other results being prepared for publication separately.

### Data analysis

Descriptive analysis of the questionnaire results was performed using SPSS version 27. Differences between the three population groups (DU, AU, RR) were assessed using the appropriate parametric or non-parametric tests (Kruskal-Wallis, or ANOVA) depending on the distribution of the variables, with further pairwise comparisons conducted (using Mann Whitney tests, or independent t-tests) where a significant difference was found on three-way testing. Differences were also assessed according to age and multimorbidity.

Thematic analysis was conducted on the interview transcripts to identify common themes in patients’ views and experiences of MDT-care and reception signposting, and to identify similarities and differences according to age, multimorbidity status and population group. Three authors (ED, SWM, KS) independently identified codes based on individual analysis of selected interview transcripts, arriving at a common coding framework through discussion. Transcripts were coded using NVivo version 12 by ED and KS. Six phases of thematic analysis (as outlined by Braun and Clarke^24^) were applied by ED and KS as follows: familiarisation with the data; generation of initial codes; searching for themes; reviewing themes; defining and naming themes; and summarising themes for a final report.

Integration and synthesis of findings from the quantitative and qualitative components of this study were conducted by KS and SWM according to Farmer and colleagues’ triangulation protocol, using a convergence coding matrix to identify areas of convergence, divergence and silence^25, 26^.

## Results

### Survey results

The characteristics of survey respondents, and comparison with non-responders, are described in our previous paper and summarised in Supplementary Table 1^18^. Survey results are summarised in Tables 1 and 2. Overall, fewer than half of respondents were aware of link workers (31%), mental health nurses (32%) or pharmacists (42%). Awareness was slightly higher for physiotherapists (58%) and much higher for nurses (86%). The AU group reported lower awareness of all MDT roles than both the DU and RR groups, except for nursing staff, where DU and AU groups were similar (p<0.01). Awareness of nursing, pharmacy and physiotherapy staff was significantly higher in RR areas than in both other groups (p<0.01), while awareness of link workers and mental health nurses were similar between DU and RR areas. Awareness of physiotherapy and pharmacy roles was also higher in patients with multimorbidity than those without (p<0.01). Patients aged 65 years or older were significantly more likely to be aware of the nurse role (p<0.05) and less likely to be aware of the link worker role than those under 65 (p<0.01).

**Table 1.**
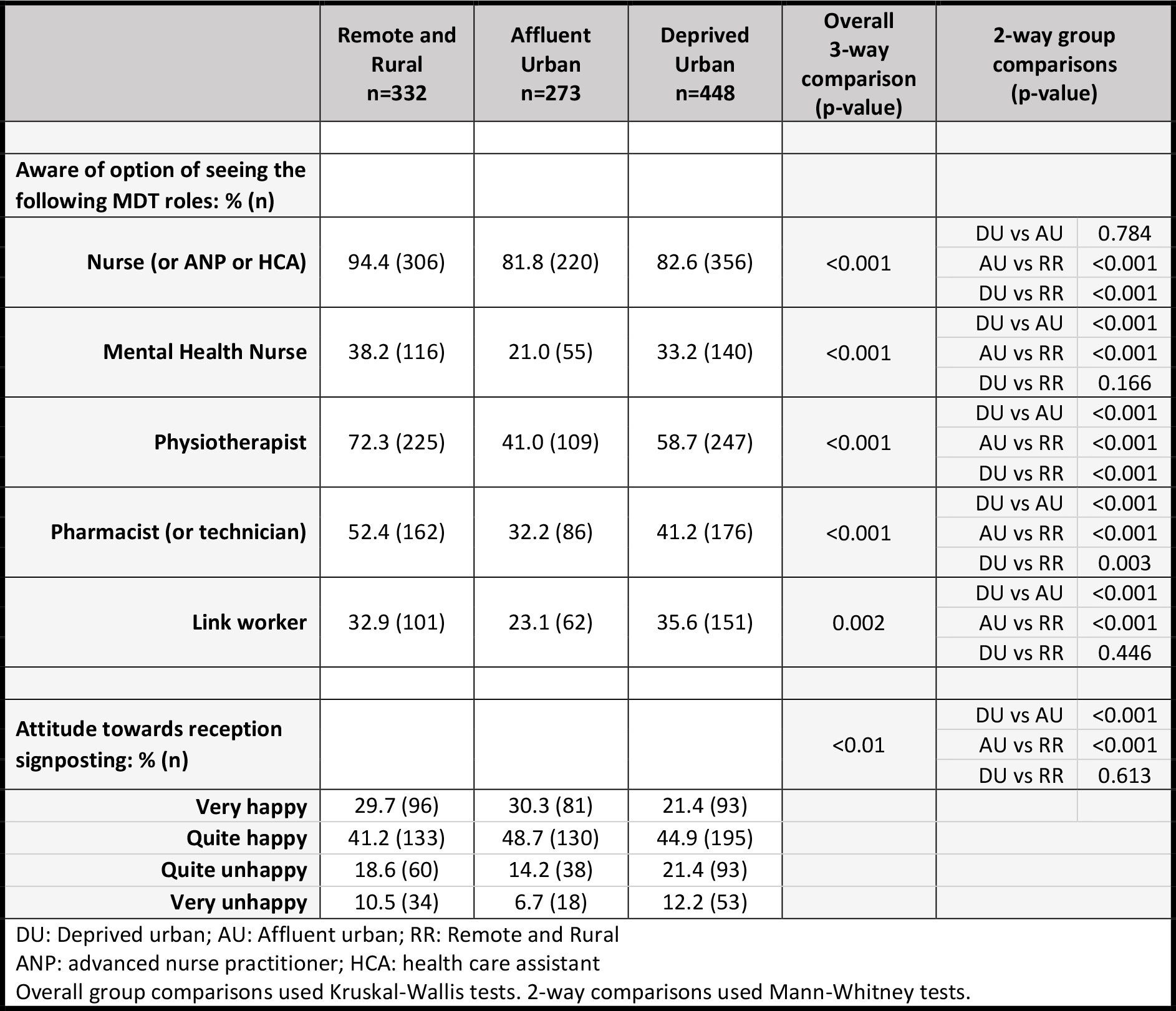
Awareness of MDT roles in participating patients from affluent urban, deprived urban, and remote and rural areas.

**Table 2.** Awareness of MDT roles comparing patients with vs without multimorbidity; and those aged 65 years and over vs those under 65.

|  | Multimorbid<br>n=727 | Non-multimorbid<br>n=322 | p-value | Aged 65<br>years and<br>over<br>n=530 | Aged under<br>65 years<br>n=498 | p-value |
| --- | --- | --- | --- | --- | --- | --- |
| Aware of option of<br>seeing the following<br>MDT roles: % (n) |  |  |  |  |  |  |
| Nurse (or ANP or<br>HCA) | 81.7 (617) | 83.8 (263) | 0.149 | 88.6 (467) | 83.4 (412) | 0.016 |
| Mental Health Nurse | 32.7 (222) | 28.6 (88) | 0.196 | 29.2 (145) | 33.6 (164) | 0.146 |
| Physiotherapist | 62.1 (430) | 49.0 (149) | <0.001 | 60.3 (307) | 55.8 (271) | 0.226 |
| Pharmacist (or<br>technician) | 44.7 (310) | 36.7 (113) | 0.017 | 43.9 (225) | 40.2 (196) | 0.007 |
| Link worker | 31.0 (213) | 32.4 (100) | 0.659 | 27.6 (140) | 35.5 (173) |  |
| Attitude towards<br>reception<br>signposting: % (n) |  |  | 0.005 |  |  | 0.439 |
| Very happy | 24.5 (173) | 30.9 (97) |  | 25.0 (132) | 27.8 (137) |  |
| Quite happy | 44.7 (316) | 44.9 (141) |  | 45.7 (241) | 44.0 (217) |  |
| Quite unhappy | 19.1 (135) | 17.8 (56) |  | 19.0 (100) | 18.1 (89) |  |
| Very unhappy | 11.7 (83) | 6.4 (20) |  | 10.2 (54) | 10.1 (50) |  |

ANP: advanced nurse practitioner; HCA: health care assistant

Attitudes towards reception signposting (Table 1) were positive on the whole, with 71% of respondents being very (26%) or quite (45%) happy with this system. There was no significant difference between AU and RR areas, but there were significantly more negative views in the DU group (34% very or quite unhappy vs 21% AU vs 29% RR; p<0.05). Acceptability of reception signposting (Table 2) was also significantly lower for patients with multimorbidity (31% quite or very unhappy vs 24% in those without; p<0.05).

### Interview results

The demographic details and multimorbidity status of interviewees across the three health boards are given in Supplementary Table 2.

#### Theme 1: Attitudes towards MDT expansion

The majority (n=22) of 30 patients interviewed were open towards the idea of receiving care from MDT staff for appropriate issues. Patients reported that this brought benefits including improved access to appointments, and in some situations, greater expertise.

> *P9. (40-49 F, SIMD 2)*
>
> *Having access to different services for people that might be more in line with what your complaint is I think can only be a good thing. If I’m being told there’s [another professional] that would better suit my needs, that I then I would go with that*.
>
> *P10. (20-29 M, SIMD 4)*
>
> *If [seeing another professional] would still get to the root of the problem and potentially negate the long waiting times of waiting for a GP, then, yes, certainly that’s something I’d be interested in. [For my condition], I would be totally happy to see a qualified physio, because that’s going to be better for me than seeing a GP*.

Some patients (n=8) expressed reservations about MDT-care, and a preference for seeing a GP as the first point of contact. The value of the doctor-patient relationship and holistic nature of a GP assessment were cited as reasons for this.

> *P21. (80-89 F, SIMD 4)*
>
> *On one side, [seeing other professionals] helps the GP. But on the other side, I think when you see the GP, they actually know you, they see how you are and sort of pick up on not just your ailment but also your mental health. [The GP] could be seeing them for, say, a migraine, but there might be a drunken husband or a runaway child. [If they] know a wee bit about the history of the family they’ve got a bit more knowledge. When [care] is dotted around different people all seeing you, some of that can get lost, you know? Some of what else is happening in the background can get totally lost. That is just one of the things that worries me [about these changes]*.

By contrast, MDT care was felt to be most appropriate for simple, focussed, task-orientated encounters.

> *P22. (60-69 F, SIMD 4)*
>
> *My consultation [with the nurse practitioner] was fine because it wasn’t for anything that I felt a doctor should be consulted for. It was to do with a fungal infection on my toenail. So, that’s fine. And I don’t expect to see a doctor for blood tests and things like that… [But] when my husband was ill, we saw a nurse practitioner, [and] personally I think I would have been happier to see a doctor because of the state of his illness*.

The importance of being able to see a GP when necessary for worrying issues was also widely expressed, including amongst patients who were more open to MDT-care.

> *P18. (70-79 F, SIMD 10)*
>
> *I still would rather value a GP’s opinion than any of the other healthcare professionals. It does depend very much on what your query is but if you’re feeling really ill I think you need to have a GP examining you. I think you should be listened to carefully about your symptoms before you’re referred to somebody other than the GP*.

#### Theme 2: Views on reception signposting

While many patients were comfortable being signposted towards MDT-care by receptionists, others were uncomfortable with this system. However, these reservations related more to the interaction with receptionists (including concerns about privacy) rather than concerns about MDT-care per se. Patients who expressed concerns about the reception signposting were more likely to live in DU or RR areas.

> *P22. (60-69 F, SIMD 4)*
>
> *I don’t think the onus should be put on the receptionist. It never used to happen. I’m a bit hesitant [about it]. I don’t like discussing [personal issues] with the receptionist. I don’t know why they should be asking. If you’re making an appointment with the GP, then you obviously feel you need the GP, you know? I don’t make an appointment for the sake of it. [Reception signposting] kind of undermines your request to see a GP*.
>
> *P26. (60-69 M, SIMD 3)*
>
> *It’s not their place. They are just the receptionist. I’m not saying they are not intelligent*… *but suddenly they have the authority to send you to a physio or refer you to a pharmacist? What authority does a receptionist act on?*

On the other hand, many patients (particularly in AU areas) had no objections to the reception signposting system and recognized its value.

> *P13. (60-69 F, SIMD 8)*
>
> *It’s good if they know who would be the best person to help you. Sometimes, with the training that the receptionists have had, some of them are fantastic. They will say, well, actually it’s probably better if you see this person rather than that person. They are a very valuable member of the team because they know exactly where to point you for you to get the best outcome. So I’m quite happy for them [to ask]*.

#### Theme 3: Experiences of MDT-care

Almost all patients interviewed had received MDT-care and described positive experiences (n=27). Encounters with nurses (either advanced nurse practitioners or practice nurses) were most common (n=20). Physiotherapy (n=10) and pharmacy (n=7) consultations were also familiar to many patients, and were felt to be particularly beneficial in terms of expertise.

> *P26. (60-69 M, SIMD 3)*
>
> *The physio was very attentive, told you what to do and why you had to do it. They explained things a bit better [than the GP]. That’s what I felt anyway. It much easier [to arrange that appointment]. Face-to-face straight away, done and dusted. And it’s an easier experience seeing another healthcare professional, it’s a more relaxed experience. I’m not saying the GP is a bad experience, but it’s often, “what’s wrong with you? Aye, this, that, that’s fine, oh aye”. That’s it and away you go, you know what I mean? [With the physiotherapist] there was more time*.

Mental health nurse (n=2) and Link Worker (n=1) encounters were least common, occurring only in practices serving DU areas. But the value of these encounters was also evident.

> P6. (50-59 M, SIMD 1)
>
> When I started to go on this Universal Credit, because I didn’t understand it, there was someone in the practice [a Link Worker] that explained it to me and told me what I was entitled to and helped me get what I was entitled to. [That was] unbelievable. Seriously, really helpful.

### Synthesis of quantitative and qualitative results

Table 3 gives the convergence coding matrix derived from the triangulation of survey and interview results. There was a high degree of convergence on several findings. Given the sequential explanatory design of the study (with interviews occurring after the survey and exploring wider, contextualising questions), some themes covered in the interviews did not have corresponding data in the survey (coded as ‘silence’ on the matrix). There was also some divergence of findings, namely that the high awareness of pharmacy and physiotherapy roles reported by survey respondents in RR areas was not reflected in the interviews. This is likely due to the small sample size of interviewees.

**Table 3.** Triangulation matrix for the findings from the quantitative and qualitative components of the study.

| Theme | Survey findings | Interview findings | Convergence coding |
| --- | --- | --- | --- |
| Awareness of the availability of MDT appointments | High awareness of nurse appointments (86%). Intermediate awareness of pharmacy (42%) and physiotherapy (58%) roles. Low awareness of MHN (32%) and LW (31%) appointments. | Most patient had experience of nurse appointments (n=20). Physiotherapy (n=10) and pharmacy (n=7) were familiar to many. MHN (n=3) and LW (n=1) encounters were less common. | Convergence |
| Differences in MDT-role awareness between patient groups | High awareness of nurse appointments in all groups, but significantly higher in RR areas.<br>Awareness of physiotherapy and pharmacy roles was highest in RR areas.<br>Awareness of MHN and LW roles was lowest in AU areas, and similar in DU and RR areas.<br>Awareness of physiotherapy and pharmacy roles higher in patients with multimorbidity than without. | Experience of nurse encounters were common in patients across all population groups (DU, AU and RR).<br>Most physiotherapy encounters were reported by patients in DU areas. Most pharmacy encounters were reported by patients in AU areas.<br>MHN and LW encounters were only reported by patients in DU areas.<br>9 interviewees (out of 30) were non-multimorbid, of which only one had experience of pharmacy or physiotherapy care. 21 interviewees were multimorbid, of which 16 had experience of pharmacy or physiotherapy care. | Partial convergence<br>Divergence<br>*Contrary to the survey, interviewees from RR areas had comparatively little experience of physiotherapy or pharmacy care.*<br>Partial convergence<br>Convergence |
| Acceptance of reception signposting | Attitudes towards reception signposting were favourable overall (71% quite or very happy). This was significantly lower in patients in DU areas. | Most patients (n=20) were, on balance, supportive of reception signposting, but many (n=17) raised concerns. Concerns were equally particularly amongst patients from DU and RR areas. | Partial convergence |
| Attitudes towards MDT care | No data | Most interviewees were open to the role of MDT in primary care. Some expressed reservations about MDT appointments as an alternative to GP care, citing the importance of GP access and holistic, relationship-based care. | Silence<br>*Attitudes towards MDT care were only explored in qualitative interviews, so there is silence on this from the quantitative component of the study.* |
| Experiences of MDT care | No data | Experiences of MDT care were overwhelmingly positive and covered all types of MDT staff. | Silence<br>*Experiences of MDT care were only explored in qualitative interviews, so there is silence on this from the quantitative component of the study.* |
| MHN = mental health nurse; LW = link worker; AU = affluent urban; DU = deprived urban; RR = remote and rural |  |  |  |

## Discussion

### Summary

Four years since the introduction of the new Scottish GP contract, patient awareness of MDT expansion is limited, views on reception signposting mixed, though experiences of MDT-care generally positive. Improved access and added expertise were perceived benefits. However, patients still want to see a known GP when they feel it is important, and report this as being challenging especially in deprived areas. Some voiced concerns that MDT-expansion came at the expense of holistic, relationship-based GP care.

There were differences in patients’ awareness of MDT roles and acceptance of reception signposting across geographic and socioeconomic groups, as well as by age and multimorbidity. Patients in DU areas were less comfortable with reception signposting, but the reasons for this relate more to concerns about privacy, rather than attitudes towards MDT-care. Pharmacy and physiotherapy roles were most familiar to patients with multimorbidity, while awareness of mental health nurses and link workers was lowest in AU areas.

### Comparison with existing literature

The differences found in MDT-role awareness between DU, AU and RR areas likely reflects differing exposure to MDT-care according to the health needs of these groups. For instance, patients in AU areas (where MDT awareness was lowest) have lower levels of multimorbidity, lower frequency of GP attendance and better general health than patients in DU and RR areas^18, 22, 27, 28^. However, the present survey found significantly higher awareness of mental health nurses in RR than AU areas, despite the fact that mental-physical multimorbidity and levels of anxiety and depression have been found to be similar between AU and RR areas^18^. The reasons for this discrepancy are not clear.

The results of this study – in particular low awareness of most MDT roles – should be considered in the context of the implementation of the 2018 contract. Local variations in the implementation of the GP contract have been reported, including the provision of MDT services through ‘hubs’ rather than individual GP practices^16, 29^. Moreover, our recent qualitative evaluations of the views of GPs, MDT staff, cluster quality leads and senior stakeholders highlighted the range of challenges affecting the expansion and integration of primary care MDTs in Scotland^16, 17^.

Both the levels awareness of MDT roles and the acceptability of reception signposting found in this study are very similar to those demonstrated in a recent Scottish Government survey with a nationally representative sample of over 1000 patients^30^. This also showed that patients view the GP as the first point of contact and prefer signposting by a GP, results which echo the findings of our qualitative interviews. Additionally, the Scottish Government survey found high levels of trust in the advice of nurses, physiotherapists and pharmacists in primary care (76%, 66% and 65% respectively; compared with 75% with complete trust in GPs and 52% in mental health nurses)^30^. These results fit with our interview findings of positive experiences of care for these three MDT roles in particular.

A number of reviews have highlighted the paucity of evidence evaluating patients’ views and experiences of MDT care^8, 13, 19^. However, our findings of broadly positive patient experiences are consistent with a recent integrative review of 48 international studies^20^. Interestingly, and in close alignment with our study, as well as finding overall positive patient experiences of MDT-care, this review highlighted concerns that increased access to MDT-care did not necessarily increase care quality, and that MDT-care was “inconsistently holistic”^20^.

Evidence on patients’ views regarding reception signposting is also lacking, although a recent study of staff views reported concerns over the lack of clarity over receptionists’ role and remit, and the need for appropriate training and development^31^. These findings echo the reservations expressed by patients in our study. While this paper focusses on the views of patients with respect to MDT expansion (to which reception signposting is an adjacent issue) a future paper is under preparation presenting a more comprehensive summary of patients’ views and experiences of primary care in Scotland.

### Strengths and limitations

The strengths of this study include its use of mixed-methodology to explore and evaluate patient views, the relatively large survey sample and the inclusion of three populations of interest. The strength of the survey findings are limited by the low response rate of 17% (12% DU vs 27% AU vs 20% RR) although this is not dissimilar to that seen in Scottish Government patient surveys^32^. Of note the DU group, which, like in other surveys, had a much lower response rate, was the biggest group in our sample, comprising 58% of distributed surveys. The response rate may also have been limited by the use of a postal survey, rather than in-person data collection at the GP practice, a method which was dictated by the pandemic. Additionally, the funding constraints of the study meant that postal reminders were not possible.

Survey responders differed from non-responders in terms of age in all groups (AU, DU and RR). In the DU group, responders were also significantly less deprived than non-responders (Supplementary Table S1). While this affects the generalisability of the survey findings, it may in fact mean that the differences found between the DU and AU groups are an underestimate, as those responding were the “least deprived of the deprived”.

With respect to the qualitative findings, the reliance on telephone rather than face-to-face interviews as a result of the pandemic may have affected the quality of evidence obtained, given the absence of non-verbal communication, although research suggests there is little difference in quality between these modes of interview^33^. The recruitment of interviewees from a volunteer subset of questionnaire respondents creates a risk of bias since this method may give prominence to those with strong views. However, while our findings do not claim to be generalizable, the number of interviews conducted, the purposive sampling method and the saturation of data suggest that our findings give a valuable representation of the range of views held by patients across geographic and socioeconomic groups and, in particular, older patients with multimorbidity.

### Implications for practice, policy and research

MDT expansion is a core feature of not only the Scottish GP contract, but of primary care reforms in healthcare systems globally^13^, and the views and experiences of patients with respect to this are under-explored^8, 13, 19^. Consequently, this study provides an important insight for all primary care systems undergoing change.

The implementation of the new GP contract in Scotland has clearly been affected by the pandemic, and other challenges to MDT expansion have been identified^16, 17^, but further work is needed to assess variations in the availability of primary care MDT services across Scotland, as this has implications for interpreting differences in patient views on these reforms. Our findings, however, suggest that despite low awareness of MDT-care, it has broad acceptance and leads to positive patient experiences. Importantly, these findings are qualified by patients’ reservations about reception signposting (which are stronger in DU areas), and overriding preference for holistic, relationship-based GP-care.

One of the key objectives of MDT expansion in Scotland was to enable GPs to focus more time on patients with complex needs. Our recent qualitative studies suggest this is not happening^16, 17^, even though evidence suggest it would be both cost-effective and beneficial^34, 35^. The preference of patients for first contact GP care suggested in this study, and supported elsewhere^30^, represents one barrier to this. Further work is needed to ensure the contract delivers on its aim of improving care for people in Scotland.

## Data Availability

All data produced in the present study are available upon reasonable request to the authors

---

**Supplementary Table 1.** Sociodemographic characteristics of participating patients in affluent urban, deprived urban, and remote and rural areas.

|  | Remote and Rural<br>n=332 | Affluent Urban<br>n=273 | Deprived Urban<br>n=448 | Overall group comparison<br>(p-value) | 2-way group comparisons<br>(p-value) |  |
| --- | --- | --- | --- | --- | --- | --- |
| **Sex: % (n)** |  |  |  | 0.491 |  |  |
| Male | 39.8 (128) | 42.8 (113) | 37.9 (163) |  |  |  |
| Female | 60.2 (194) | 56.4 (149) | 61.9 (266) |  |  |  |
| Other | 0 (0) | 0.8 (2) | 0.2 (1) |  |  |  |
| **Age: mean (SD)** | 65.8 (14.5) | 62.0 (17.6) | 60.9 (14.9) | <0.001 | DU vs AU | 0.001 |
|  |  |  |  |  | AU vs RR | <0.001 |
|  |  |  |  |  | DU vs RR | 0.405 |
| **Age group: % (n)** |  |  |  | <0.001 | DU vs AU | 0.217 |
|  |  |  |  |  | AU vs RR | 0.032 |
|  |  |  |  |  | DU vs RR | <0.001 |
| <45 years | 9.3 (30) | 17.5 (47) | 14.2 (62) |  |  |  |
| 45-64 years | 31.5 (102) | 29.5 (79) | 40.8 (178) |  |  |  |
| 65+ years | 59.3 (192) | 53.0 (142) | 45.0 (196) |  |  |  |
| **SIMD decile: % (n)** |  |  |  | <0.001 | DU vs AU | <0.001 |
|  |  |  |  |  | AU vs RR | <0.001 |
|  |  |  |  |  | DU vs RR | <0.001 |
| Most deprived 1 | 0 (0) | 0 (0) | 19.5 (87) |  |  |  |
| 2 | 0.3 (1) | 0 (0) | 23.5 (105) |  |  |  |
| 3 | 4.2 (14) | 1.5 (4) | 13.0 (58) |  |  |  |
| 4 | 32.2 (107) | 5.5 (15) | 12.1 (54) |  |  |  |
| 5 | 20.2 (67) | 2.2 (6) | 6.7 (30) |  |  |  |
| 6 | 22.6 (75) | 0.4 (1) | 8.1 (36) |  |  |  |
| 7 | 15.1 (50) | 3.3 (9) | 4.9 (22) |  |  |  |
| 8 | 2.1 (7) | 6.2 (17) | 2.9 (13) |  |  |  |
| 9 | 3.3 (11) | 15.0 (41) | 6.5 (29) |  |  |  |
| Least deprived 10 | 0 (0) | 65.9 (180) | 2.9 (13) |  |  |  |
| **Job status: % (n)** |  |  |  | 0.007 | DU vs AU | 0.104 |
|  |  |  |  |  | AU vs RR | 0.321 |
|  |  |  |  |  | DU vs RR | 0.002 |
| Employed | 34.5 (112) | 40.1 (107) | 39.8 (174) |  |  |  |
| Retired | 58.8 (191) | 52.1 (139) | 41.2 (180) |  |  |  |
| Unemployed, looking | 0.3 (1) | 0.7 (2) | 2.1 (9) |  |  |  |
| Unemployed, unable | 5.2 (17) | 1.9 (5) | 14.4 (63) |  |  |  |
| In education | 0.3 (1) | 2.2 (6) | 0.5 (2) |  |  |  |
| **Living arrangement: % (n)** |  |  |  | 0.120 |  |  |
| Alone | 24.6 (80) | 24.0 (64) | 35.2 (153) |  |  |  |
| With partner | 68.6 (223) | 69.7 (186) | 53.3 (232) |  |  |  |
| With someone else | 6.8 (22) | 6.4 (17) | 11.5 (50) |  |  |  |
| **Time elapsed since consultation: % (n)** |  |  |  | 0.803 |  |  |
| Less than 1 week | 9.1 (30) | 8.9 (24) | 8.7 (38) |  |  |  |
| 1-2 weeks | 18.2 (18.2) | 17.1 (46) | 16.7 (73) |  |  |  |
| 2-3 weeks | 28.3 (93) | 34.6 (93) | 33.4 (146) |  |  |  |
| 4 or more weeks | 44.4 (146) | 39.4 (106) | 41.2 (180) |  |  |  |

DU: Deprived urban; AU: Affluent urban; RR: Remote and Rural
Overall group comparisons used Kruskal-Wallis tests, except for Age and SIMD where ANOVA was used.
2-way comparisons used Mann-Whitney tests, except for Age and SIMD, which used independent t-tests.

**Supplementary Table 2.** Characteristics of interviewees.

| **Health Board 1** | **Practice demographic** | **Age group** | **Gender** | **SIMD** | **No. of LTC** |
| --- | --- | --- | --- | --- | --- |
| P1 | Deprived urban | 30-39 | Female | 4 | 4 |
| P2 | Deprived urban | 70-79 | Female | 2 | 5 |
| P3 | Deprived urban | 60-69 | Male | 1 | 4 |
| P4 | Deprived urban | 60-69 | Female | 2 | 8 |
| P5 | Deprived urban | 70-79 | Male | 3 | 2 |
| P6 | Deprived urban | 50-59 | Male | 1 | 2 |
| P7 | Deprived urban | 60-69 | Male | 2 | 4 |
| P8 | Deprived urban | 60-69 | Female | 5 | 1 |
| P9 | Deprived urban | 40-49 | Female | 2 | 1 |
| P10 | Deprived urban | 20-49 | Male | 4 | 2 |
| **Health Board 2** | **Practice demographic** | **Age group** | **Gender** | **SIMD** | **No. of LTC** |
| P11 | Affluent urban | 70-79 | Male | 9 | 6 |
| P12 | Deprived urban* | 30-39 | Female | 3 | 4 |
| P13 | Deprived urban* | 60-69 | Female | 8 | 5 |
| P14 | Affluent urban | 70-79 | Male | 10 | 4 |
| P15 | Affluent urban | 70-79 | Male | 10 | 4 |
| P16 | Deprived urban* | 50-59 | Female | 7 | 4 |
| P17 | Deprived urban* | 60-69 | Female | 9 | 6 |
| P18 | Affluent urban | 70-79 | Female | 10 | 1 |
| P19 | Affluent urban | 30-39 | Male | 10 | 1 |
| P20 | Affluent urban | 70-79 | Male | 10 | 1 |
| **Health Board 3** | **Practice demographic** | **Age group** | **Gender** | **SIMD** | **No. of LTC** |
| P21 | Remote and rural | 80-89 | Female | 4 | 9 |
| P22 | Remote and rural | 60-69 | Female | 4 | 4 |
| P23 | Remote and rural | 70-79 | Female | 6 | 7 |
| P24 | Remote and rural | 70-79 | Male | 5 | 3 |
| P25 | Remote and rural | 50-59 | Female | 5 | 4 |
| P26 | Remote and rural | 60-69 | Male | 3 | 6 |
| P27 | Remote and rural | 60-69 | Male | 5 | 2 |
| P28 | Remote and rural | 60-69 | Male | 4 | 1 |
| P29 | Remote and rural | 20-29 | Female | 5 | 1 |
| P30 | Remote and rural | 70-79 | Male | 4 | 0 |
| SIMD = Scottish index of multiple deprivation (1=most deprived; 10=least deprived) |  |  |  |  |  |
| LTC= Long term conditions |  |  |  |  |  |
| *These patients were from practices in Health Board 2 serving mainly deprived urban areas, but which also included some affluent postcodes. |  |  |  |  |  |

